# Screening and treatment time in school-based caries prevention: A randomized clinical trial

**DOI:** 10.1101/2024.07.11.24310306

**Authors:** Tamarinda J. Barry Godín, Gabriel Hawthorne, Radhika Shah, Ryan Richard Ruff

**Affiliations:** Department of Epidemiology & Health Promotion, New York University College of Dentistry, New York, NY, 10010; New York University School of Global Public Health, New York, NY, 10010

## Abstract

**Background:** School-based caries prevention can increase access to dental services for underrepresented children and reduce the risk of tooth decay.

**Methods:** The CariedAway study was a longitudinal pragmatic randomized trial of silver diamine fluoride (SDF), fluoride varnish, dental sealants, and atraumatic restorations (ART) provided as part of a school caries prevention program. Using electronic health record software and reproducible procedures, we estimated the total time required to screen and treat program participants. Differences at initial treatment between interventions, provider (registered nurse and dental hygienist), dentition mix, and caries burden were determined using linear regression with cluster standard error estimation, and longitudinal effects were estimated using linear mixed effects models.

**Results:** A total of 7418 children were enrolled in the CariedAway trial, of which 7176 (97%) had viable data recorded for screening and treatment time. Overall treatment time for children receiving SDF and fluoride varnish was 283 seconds (SD=739), compared to 753 seconds (SD=2166) for children receiving dental sealants and ART. At the initial program visit, treatment time using SDF was significantly shorter than sealants and ART (B = −458.8, 95% CI = −650.1, −266.8) and treatment time decreased with each subsequent observation (B = −51.9, 95% CI = −68.4, −35.4). Treatment time significantly increased as the number of carious teeth per child increased, and there were no differences in treatment time using SDF between registered nurses and dental hygienists.

**Conclusions:** The sustainability of school-based caries prevention can be supported by robust data on program logistics and treatment time. These results can be leveraged by future school-based sealant and SDF programs to estimate the total reach and effectiveness of intended treatments.

## Introduction

Dental caries (tooth decay) is a global public health concern affecting billions of individuals across the lifespan [1]. Untreated caries poses significant health risks to school-age children including pain and infection, and can have adverse effects on psychosocial functioning [2], academic performance and school attendance [3], and quality of life [4]. Low-income and minority children experience profound inequities in both oral health outcomes and access to dental care [5]. In the United States, the prevalence of untreated caries in primary dentition amongst children from families with an income less than 100% of the federal poverty level is nearly three times that of other children, and the prevalence in black and Mexican American children is twice that of white children [6].

School-based caries prevention can effectively increase access to care and reduce oral health inequities [7]. For example, school sealant programs can protect against caries in vulnerable children [8] and are officially recommended by both the CDC and the Community Preventive Services Task Force [9, 10]. In addition to dental sealants [8, 11], other clinical approaches for school-based caries prevention include fluoride rinses [12], fluoride varnish [13], atraumatic restorations (ART) [14], and silver diamine fluoride (SDF) [15, 16]. The sustainability of these programs depends on a multitude of factors including clinical- and cost-effectiveness, convenience, flexibility in workforce requirements, community engagement, and program participation. While the clinical and economic impact of school caries prevention is well-established [10, 11], little data is available on the implementation of school-based care.

*CariedAway* was a pragmatic clinical trial of minimally invasive approaches to prevent and control caries in schools [17], treating children with fluoride varnish, silver diamine fluoride, dental sealants, and atraumatic restorations. Study procedures in CariedAway used reproducible clinical protocols for surface-level tooth assessment and treatment, as well as electronic data entry for robust measures of time. Our objective was to assess screening and treatment time for school-based caries prevention to better inform future program development and implementation.

## Methods

CariedAway is a registered study at www.clinicaltrials.gov (#NCT03442309) and is reported according to CONSORT recommendations for randomized trials. The study received ethical approval from the New York University School of Medicine IRB (#i17-00578). A study protocol is available [17].

### Design and Participants

The CariedAway study was a longitudinal, pragmatic, cluster-randomized, non-inferiority trial conducted in primary school children in New York City from February 1, 2019, through June 1, 2023. The original objectives of CariedAway were to compare minimally invasive dental procedures, provided in a school-based program, in the two year arrest rate and four year prevention rate of dental caries. All schools participating in the study were required to have a student population of at least 50% black and/or Hispanic/Latino and at least 80% receiving free and reduced lunch, which is a proxy indicator for low socio-economic status. Inclusion criteria for children in participating schools included parental informed consent and child assent.

### Randomization

Schools included in CariedAway were block randomized at the school level to each provided treatment using a random number generator. All participating subjects in schools received the same treatment.

### Standardization and Calibration

Approximately 70 hours of didactic and practical training, including clinical presentations and hands-on application of preventive techniques utilizing typodonts, was conducted with clinical staff, followed by digital assessments to evaluate major concepts. Feedback was provided, and peer learning sessions were conducted to reinforce understanding. Examiner training sessions involved live subjects presenting with carious lesions of varying severity. Findings were systematically reviewed by a senior examiner (licensed dentist), with repeated assessments conducted until inter-examiner agreement was achieved. This standardization process, performed at least annually for all clinical personnel and in a one-on-one capacity for new personnel, facilitated a systematic approach to dental screening and treatment provision.

### Clinical procedures: Screening

A standardized clinical protocol was employed for surface-level tooth assessment utilizing the International Caries Detection and Assessment System’s (ICDAS) adapted criteria in epidemiology and clinical research settings [18]. Any lesions with an ICDAS score of 5 or 6 was assessed as untreated caries. A visual-tactile dental screening was performed utilizing a dental mirror and explorer, frontal light source, and an electronic data entry device (tablet computer). Dental screenings were conducted in two rounds, with examiners systematically inspecting teeth in the arch starting from the upper right and lower left quadrants of the mouth, proceeding mesially and crossing the midline. Data were recorded in real-time by a dedicated recorder with the examiner providing tooth names and surface-level findings. Quadrant completion was then marked to ensure consistency between examiner findings and data entry.

### Clinical procedures: Treatment

Treatments were administered by either registered dental hygienists or registered nurses under the availability of patient-specific standing orders signed by the supervising dentist. Those randomly assigned to the experimental group received an application of 38% silver diamine fluoride (Elevate Oral Care Advantage Arrest 38%, 2.24 F-ion mg/dose) on posterior, asymptomatic, cavitated lesions and the pits and fissures of all sound bicuspids and molars. After cleaning and drying affected tooth surfaces, a microbrush was used to transfer SDF to individual teeth for a minimum of 30 seconds, followed by 60 seconds drying time. Subjects assigned to the active control received glass ionomer sealants (GC Fuji IX, GC America) on the pits and fissures of all sound bicuspids and molars, and the placement of atraumatic restorations on all frank, asymptomatic, cavitated lesions. Participants in both treatment groups then received fluoride varnish (5% NAF, Colgate PreviDent) applied to all teeth. All treatments were provided in a dedicated room in a school (e.g., an empty classroom) using a dental mirror and explorer, frontal light source, and portable dental chairs.

### Outcome derivation

The time required for each individual screening and treatment was calculated using automatically-generated time indicators produced by chairside electronic health record (EHR) software. Each EHR for study participants consisted of patient information, intra- and extra-oral findings, oral screening, treatment plan, and visit summary pages. Screening time was assessed by extracting and calculating the difference between time stamps for the first entry on the treatment planning page of the EHR and the beginning of the intra- and extra-oral findings page of the EHR. Treatment time was assessed by extracting and calculating the difference between time stamps for activation of the visit summary page of the EHR, which completes the treatment protocol, and the first entry on the treatment planning page. Full description of the specific clinical procedures corresponding to these periods of time are included in Supplementary Materials.

### Statistical Analysis

Descriptive statistics were computed for the full study sample and by treatment group for sociodemographic variables (e.g., sex, race/ethnicity) and the prevalence of untreated decay at baseline. The total raw time in seconds was then estimated for screening and treatment by select study and clinical variables, including treatment group, the number of caries present at the time of treatment, whether care was provided by a registered nurse or dental hygienist, dentition type (e.g., mixed dentition, primary teeth only, and permanent teeth only), and whether the participant had urgent needs defined as presence of pain, abscess, swelling, fistula, or visible pulpal involvement. Unadjusted and adjusted differences in treatment time at initial observation, as well as longitudinal models, were estimated using mixed effects multilevel models. Random effects were included for clustering at the school level and for observations within patients (for longitudinal models). Statistical significance was determined at p < 0.05. Analysis was performed using R v4.4.0.

## Results

A total of 7418 children were enrolled in the CariedAway trial; 3739 (50.4%) were randomized to receive SDF and 3679 (49.6%) randomized to receive dental sealants and ART (Table 1). Approximately 27% of study participants had untreated decay at baseline and 66% were of either Hispanic/Latino or black race/ethnicities. The average age at enrollment was 7.6 years (SD = 1.90).

**Table 1:** Sample demographics.

|  | **Overall** |  | **SDF** |  | **Sealants and ART** |  |
| --- | --- | --- | --- | --- | --- | --- |
|  | N | % | N | % | N | % |
| Participants (enrolled) | 7418 | 100 | 3739 | 50.4 | 3679 | 49.6 |
| Baseline decay | 1980 | 26.69 | 1016 | 27.17 | 964 | 26.2 |
| Sex (male) | 3412 | 46 | 1785 | 47.74 | 1627 | 44.24 |
| Race/Ethnicity |  |  |  |  |  |  |
| Hispanic/Latino | 3648 | 49.18 | 1766 | 47.23 | 1882 | 51.16 |
| Black | 1246 | 16.8 | 650 | 17.38 | 596 | 16.2 |
| White | 153 | 2.06 | 86 | 2.3 | 67 | 1.82 |
| Asian | 125 | 1.69 | 88 | 2.35 | 37 | 1.01 |
| More than one | 114 | 1.54 | 67 | 1.79 | 47 | 1.28 |
| Other | 90 | 1.21 | 56 | 1.5 | 34 | 0.92 |
| Unreported | 2042 | 27.53 | 1026 | 27.44 | 1016 | 27.62 |
| Age at baseline | 7.6 | 1.90 (SD) | 7.5 | 1.92 (SD) | 7.6 | 1.88 (SD) |

Of the study sample, 7176 (97 %) had viable data recorded for screening and treatment phases (Table 2). Across all participants, the average time required for screening was 170 seconds (SD=233), and the average time for treatment was 513 seconds (SD=1621). Screening time was similar in children treated with SDF (161, SD=260) or sealants and ART (179, SD=199), while treatment time was considerably higher in the sealant and ART group (752, SD=2165) compared to the SDF group (283, SD=739). Screening and treatment time both increased as the number of caries per-participant increased, and those with urgent needs had higher screening and treatment times than those that did not.

**Table 2:** Total average screening and treatment time at baseline by clinical indicators.

|  | N / % | Screening |  | Treatment |  |
| --- | --- | --- | --- | --- | --- |
|  |  | Mean | SD | Mean | SD |
| Overall | 7176 | 170.05 | 232.8 | 512.56 | 1621.18 |
| Treatment |  |  |  |  |  |
| SDF | 3667 | 161.28 | 260.64 | 282.9 | 739.41 |
| Sealant + ART | 3509 | 179.22 | 199.25 | 752.57 | 2165.98 |
| SDF |  |  |  |  |  |
| 0 caries | 2667 | 130.01 | 267.76 | 257.24 | 667.07 |
| 1-3 caries | 801 | 221.75 | 206 | 336.06 | 924.19 |
| >3 caries | 199 | 336.97 | 249.11 | 412.84 | 801.82 |
| Sealant + ART |  |  |  |  |  |
| 0 caries | 2578 | 155.09 | 165.95 | 650.17 | 1880.29 |
| 1-3 caries | 774 | 226.77 | 212.93 | 1026.18 | 2855.85 |
| >3 caries | 157 | 341.01 | 407.33 | 1085.16 | 2454.52 |
| SDF Provider (SDF group only) |  |  |  |  |  |
| Hygienist | 2890 | 149.69 | 268.83 | 282.13 | 790.5 |
| Nurse | 754 | 197.78 | 204.71 | 259.94 | 338.51 |
| Dentition type |  |  |  |  |  |
| Primary only | 992 (13.82) | 127.96 | 189.9 | 570.44 | 2243.24 |
| Early mixed | 2597(36.19) | 163.72 | 194.01 | 492.4 | 1480.44 |
| Late mixed | 3051 (42.51) | 190.55 | 281.34 | 502.16 | 1474.91 |
| Permanent | 536 (7.46) | 161.95 | 142.37 | 562.39 | 1696.71 |
| Severity |  |  |  |  |  |
| Non-urgent | 7021 (97.84%) | 165.8 | 224.99 | 510.1 | 1636.99 |
| Urgent | 155 (2.16%) | 360.7 | 424.53 | 626.4 | 532.31 |

*Note: Estimates in seconds*

In adjusted analyses (Table 3), treatment time with silver diamine fluoride was significantly lower than with sealants and ART (B = −458.8 seconds, 95% CI = −650.1, −266.8). Similarly, each additional tooth with dental caries took approximately 1.5 minutes longer for treatment (B = 80.63, 95% CI = 48.1, 113.1) across both groups. There were no significant differences by participant age, dentition type (e.g., mixed dentition, permanent only, or primary only), or when comparing SDF when applied by a nurse or dental hygienist. When restricted to participants with no dental caries (primary prevention only), treatment time with SDF remained significantly reduced (B = −369.1, 95% CI = −515.2, −222.9). Similarly in participants with any decay (caries prevention and control), treatment with SDF took 662.5 fewer seconds than sealants and ART (95% CI = −943.0, −381.4).

**Table 3:** Regression results for treatment time by clinical indicators, initial visit.

| *Variables* | Unadjusted |  |  |  | Adjusted |  |  |  |
| --- | --- | --- | --- | --- | --- | --- | --- | --- |
|  | **B** | **SE** | **95% L** | **95% U** | **B** | **SE** | **95% L** | **95% U** |
| SDF (vs sealant) | **-439.47** | 81.31 | -598.53 | -280.15 | **-458.84** | 167.46 | -650.12 | -266.82 |
| # of caries (continuous) | **73.95** | 13.69 | 47.02 | 100.64 | **80.63** | 16.59 | 48.1 | 113.09 |
| Age | -4.28 | 11.59 | -27 | 18.45 | 3.36 | 20.37 | -36.48 | 43.32 |
| Dentition type (ref: early mixed) |  |  |  |  |  |  |  |  |
| Late mixed | -8.089 | 57.25 | -91.33 | 75.25 | 0.12 | 67.74 | -132.77 | 132.67 |
| Permanent only | 75.63 | 42.49 | -73.39 | 224.63 | 87.89 | 118.75 | -145.21 | 320.16 |
| Primary only | 71.23 | 76.04 | -45.01 | 187.51 | 97.13 | 77.57 | -54.51 | 249.52 |
| Nurse (vs hygienist)* | 25.44 | 29.81 | -32.8 | 84.02 | 34.31 | 32.03 | -28.33 | 97 |
| *Treatment differences for prevention and control* |  |  |  |  |  |  |  |  |
| In children with no caries | **-369.12** | 74.65 | -515.19 | -222.88 |  |  |  |  |
| In children with caries | **-662.5** | 143.4 | -942.95 | -381.39 |  |  |  |  |

\*SDF group only

Longitudinal results (Table 4) for treatment group differences were similar to cross-sectional analyses, with treatment time for SDF application being significantly shorter than that for sealants and ART (B = −400.5, 95% CI = −443.2, −357.8). Additionally, each successive post-baseline visit was associated with an approximate 50 second reduction in treatment time (B = −51.9, 95% CI = −68.4, −35.4) while each additional tooth with caries resulted in a 58.3 second increase in time (95% CI = 42.2, 74.4). Finally, the interaction between treatment and time was significant, indicating that relative to sealants, treatment time for SDF increased with each observation (B = 77.9, 95% CI = 45.0, 110.9). This can be expected as SDF was reapplied at each visit while sealants were only applied if they were not retained.

**Table 4:**
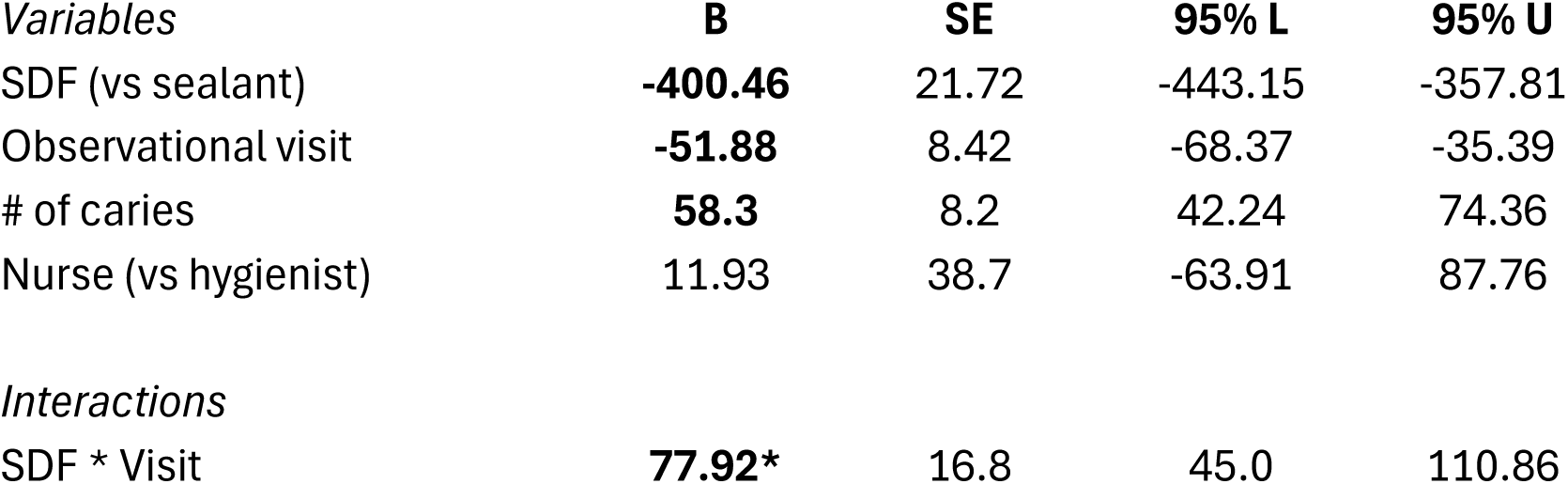
Longitudinal model results.

## Discussion

In this school-based randomized clinical trial of minimally invasive caries prevention agents, we show that the treatment time for participants receiving silver diamine fluoride and fluoride varnish was less than five minutes per child, significantly shorter than treatment with dental sealants and atraumatic restorations. We estimate that the total average time required for both screening and treatment across all children receiving SDF and fluoride varnish to be approximately seven minutes per child, compared to fifteen minutes for sealants and atraumatic restorations. These differences are consistent whether interventions were provided as primary prevention only or for both caries prevention and control. Longitudinally, treatment time decreased by approximately one minute for each post-baseline observation, while each additional tooth with caries was associated with a 1.5 minute increase in treatment time. Given that SDF is comparable to sealants and ART in preventing caries in school-based dental programs [15, 16], we conclude that SDF application is more efficient and our results highlight its potential to increase access to preventive dental care by serving a larger number of students within a given timeframe.

Silver diamine fluoride is a non-invasive, cost-effective, topical fluoride treatment recognized for its ease of application and tolerability [19]. The American Academy of Pediatric Dentistry (AAPD) chairside guide for SDF recommends at least one minute of application time and three minutes for drying per treatment [19], while the clinical practice guidelines for the use of pit and fissure sealants published by the American Dental Association (ADA) and AAPD makes no mention of application time, likely due to significant variations in material selection (e.g., auto-polymerized versus light-cured or resin-based versus glass ionomer) and application approach. For example, the National Maternal and Child Oral Health Resource Center notes that effective isolation of half of the mouth at one time will reduce sealant application time and increase efficiency [20]. In outpatient clinics, treatment time with SDF can range from approximately 50% to 80% faster than atraumatic restoration [21, 22], but may take longer when dental dams are used [23]. Additionally, these estimates may not reflect the real-world needs of school-based care, which can include working in a classroom environment, using mobile equipment, and coordinating multiple children in preparation for treatment. Prior studies of mobile clinics providing dental sealants to schools estimate an application time of approximately nine minutes for initial treatment and two minutes for repeated application [24].

School-based programs are attractive as they can increase access to dental services for a substantial number of children, particularly those at high risk for dental caries [10]. However, large schools, diverse schools, and schools serving low-income children with health disparities may require more time to treat children due to increased administrative and coordination efforts and the complex needs of the student population. Specifically, young children, children who have not previously received dental services, and children with extensive dental needs are at increased likelihood of dental anxiety and may therefore exhibit behaviors that require specialized management techniques, adding time to the treatment process [25, 26]. However, in addition to overall clinical effectiveness, research suggests that nurses are as effective as dental hygienists in using silver diamine fluoride in school-based caries prevention [27]. This may further imply that school nurses, who are likely more familiar with the students in their schools, may be particularly adept at navigating individual patient needs and further expand the reach of school dental health.

The CariedAway study included a Patient and Stakeholder Engagement Board that contributed to trial design and implementation, and included representatives from schools, local health organizations, and parents. During study development, notable concerns were raised regarding potential disruption to school schedules and student time spent away from class. Prioritizing interventions that are efficient in terms of time and resources can minimize the duration of student absences from academic programming while promoting a positive treatment experience, increasing acceptance among students, their families, teachers, and staff. Regardless of the differences observed in this analysis, school-based prevention with either sealants/ART or SDF would be preferrable to no care, as oral health issues are the leading contributor to missed school, accounting for over 30 million hours of lost school-time per year [28].

Our results indicate that as a school-based approach to caries prevention and control, silver diamine fluoride requires less time compared to dental sealants and ART, potentially leading to increased efficiency in school dental health. This efficiency can further optimize resources and streamline program implementation, which may also translate to cost savings in terms of personnel hours, materials, and overall program costs. School caries prevention using SDF can therefore lead to more effective, accessible, and sustainable oral health initiatives in school settings.

## Author Contributors

*Concept and design:* RRR

*Acquisition, analysis, or interpretation of the data:* All authors

*Drafting of the Manuscript*: TBG, RRR

*Critical review of manuscript*: All authors

*Statistical Analysis*: GH, RS, RRR

*Obtained funding*: RRR

*Administrative, technical, or material support*: TBG

*Supervision:* RRR

## Data Sharing

**Data available:** Yes

**Data types:** Data dictionary

**How to access data:** Data dictionaries will be available to interested researchers upon request to the authors

**When available:** beginning date: 12-01-2025

**Supporting Documents**

**Document types:** Informed consent form

**How to access documents:** Informed consent forms will be available to interested researchers upon request to the authors

**When available:** beginning date: 12-01-2025

**Additional Information**

**Who can access the data:** Interested researchers upon request to the authors

**Types of analyses:** For any purpose

**Mechanisms of data availability:** After approval of a proposal and a signed data access agreement.

## Data Availability

Data available: Yes
Data types: Data dictionary
How to access data: Data dictionaries will be available to interested researchers upon request to the authors
When available: beginning date: 12-01-2025
Supporting Documents
Document types: Informed consent form
How to access documents: Informed consent forms will be available to interested researchers upon request to the authors
When available: beginning date: 12-01-2025
Additional Information
Who can access the data: Interested researchers upon request to the authors
Types of analyses: For any purpose
Mechanisms of data availability: After approval of a proposal and a signed data access agreement.

## Supplementary Material

**S.1.** Screening and treatment procedures included in time estimates

1. Start New Exam and Treatment

a. Review patient identifiers

i. First name
ii. Last name
iii. Date of Birth
b. Review patient demographics

i. Current grade
ii. Race and ethnicity
iii. Contraindication of care
iv. Confirm changes in general health since the last appointment, as applicable
v. Confirm allergies to any medications, foods, or latex
c. Attain oral assent
2. Pathology

a. Assess the patient for:

i. Evidence of dry mouth
ii. Visible plaque or gingival inflammation
iii. Non-oral injury, bruise, breaks or burns
iv. Oral soft tissue lesions
v. Oral hygiene status
b. Author specific Take Home Form notes, as necessary and applicable
3. Screening/Exam Data

a. Beginning from the upper right, the clinician will begin the exam by identifying which teeth are present and missing
b. Record tooth-level information:

i. The clinician will identify whether the tooth has caries, arrested decay, sealant, filling, and/or crown present at the surface level (i.e., mesial, occlusal, distal, buccal, lingual)
ii. The clinician will check teeth for urgent/emergent needs at the tooth-level, including the presence of pulpal exposure, fistula (abscess), swelling, or pain
c. The clinician will repeat this process until all teeth have been screened and recorded
4. Treatment Data

a. Clinicians will treatment plan teeth designated to receive treatment according to either SDF or sealant/ART protocols determined by the preventive care assignment for the students attending a given school

i. Silver diamine fluoride
ii. Pit and fissure sealants and interim therapeutic restorations
5. Visit Summary

a. Verify the completion of all relevant procedures
b. Submit Additional Visit Notes as necessary and applicable:

i. Description of specific conditions
ii. Unanticipated problems that may have occurred
c. Have the examiner/screener sign and date the electronic health record
d. Have the treatment provider sign and date the electronic health record
6. Take-Home Form

a. Review Take Home Form for accuracy
b. Print

---

**Figure 1:**
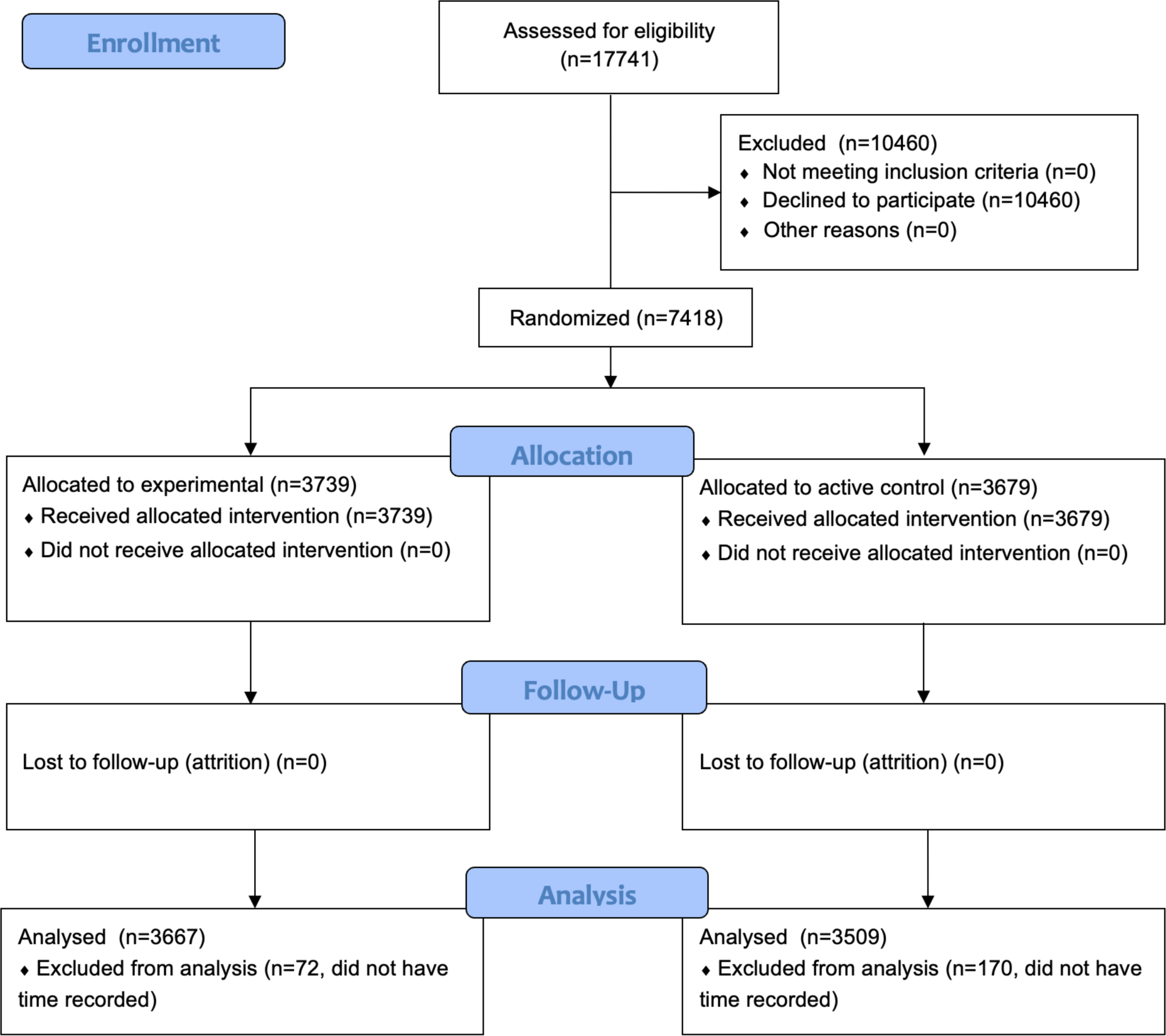
Trial enrollment flowchart.

